# Characteristics and outcomes of COVID-19 patients during B.1.1.529 (Omicron) dominance compared to B.1.617.2 (Delta) in 89 German hospitals

**DOI:** 10.1101/2022.04.09.22273420

**Authors:** Johannes Leiner, Vincent Pellissier, Sven Hohenstein, Sebastian König, Ekkehard Schuler, Robert Möller, Irit Nachtigall, Marzia Bonsignore, Gerhard Hindricks, Ralf Kuhlen, Andreas Bollmann

## Abstract

**Background:** The SARS-CoV-2 variant of concern B.1.1.529 (Omicron) was first described in November 2021 and soon became the dominant variant worldwide. Existing data suggests a reduced disease severity in comparison to B.1.617.2 (Delta). Differences in characteristics and in-hospital outcomes of patients with COVID-19 in Germany during the Omicron period compared to Delta are not thoroughly studied. Surveillance for severe acute respiratory infections (SARI) represents an integral part of infectious disease control in Germany.

**Methods:** Administrative data from 89 German Helios hospitals was retrospectively analysed. Laboratory-confirmed SARS-CoV-2 infections were identified by ICD-10-code U07.1 and SARI cases by ICD-10-codes J09-J22. COVID-19 cases were stratified by concomitant SARI. A nine-week observational period between December 6, 2021 and February 6, 2022 was defined and divided into three phases with respect to the dominating virus variant (Delta, Delta to Omicron transition, Omicron). Regression analyses adjusted for age, gender and Elixhauser comorbidities were applied to assess in-hospital patient outcomes.

**Results:** A total cohort of 4,494 inpatients was analysed. Patients in the Omicron dominance period were younger (mean age 61.6 vs. 47.8; p<0.01), more likely to be female (54.7% vs. 47.5%; p<0.01) and characterized by a lower comorbidity burden (mean Elixhauser comorbidity index 8.2 vs. 5.4; p<0.01). Comparing Delta and Omicron periods, patients were at significantly lower risk for intensive care treatment (adjusted odds ratio 0.64 [0.51-0.8]; p<0.001), mechanical ventilation (adjusted odds ratio 0.38 [0.28-0.51]; p<0.001), and in-hospital mortality (adjusted odds ratio 0.42 [0.32-0.56]; p<0.001). This also applied to the separate COVID-SARI group. During the Delta to Omicron transition, case numbers of COVID-19 without SARI exceeded COVID-SARI.

**Conclusion:** Patient characteristics and outcomes differ during the Omicron dominance period as compared to Delta suggesting a reduced disease severity with Omicron infections. SARI surveillance might play a crucial role in assessing disease severity of future SARS-CoV-2 variants.

## Introduction

Various variants of SARS-CoV-2 continue to emerge during the COVID-19 pandemic. The European Centre for Disease Prevention and Control (ECDC) and the World Health Organization (WHO) currently list several virus lineages under close monitoring including the circulating variants of concern (VoC) B.1.617.2 (Delta) and B.1.1.529 (Omicron) ^1 2^. First described in South Africa and reported to the WHO on November 24, 2021 ^3^, Omicron has become the predominant SARS-CoV-2 variant worldwide throughout the past two months, with a share of sequences in European countries of almost 100% ^4^. Several sub-lineages of Omicron exist with BA.1 and BA.2 being the most common ones ^5^. Numbers of BA.2 increased rapidly worldwide as of January 2022 ^6^ with a proportion of up to 72% among Omicron sequences recently reported for Germany by the federal government agency Robert-Koch-Institute (RKI)^7^.

Although Omicron carries several previously observed mutations, its genome markedly differs from those of other VoC which leads to significant differences in viral behaviour in comparison to Delta with respect to transmissibility and immune evasion ^8-10^. However, early data from South Africa suggested a reduced disease severity in adults with Omicron infections ^11^. In accordance therewith, reduced odds for hospitalisation or death were reported for patient cohorts in Norway ^12^ Canada ^13^ and the US ^14^. Most recently, Nyberg *et al*. presented data of over 1.5 million laboratory-confirmed COVID-19 cases in the United Kingdom and found a substantially decreased risk for hard outcomes like hospital admission or death in patients infected with Omicron as compared to Delta ^15^. The findings were consistent even when only unvaccinated individuals were analysed suggesting a “real” reduction in viral pathogenicity.

So far, outcome data of hospitalized patients in Germany infected during the Omicron dominance period is scarce. Weekly reports with descriptive data on incidence, hospitalizations and mortality are presented by the RKI ^16^, however, structured statistical analyses are lacking. The aim of this study was to evaluate characteristics and outcomes of hospitalized patients with COVID-19 in Germany, estimate the disease severity of Omicron and investigate temporal trends during the Delta to Omicron transition phase. To serve this purpose, we assessed the frequency of severe acute respiratory infections (SARI) in COVID-19 patients.

## Methods

We conducted a retrospective claims data-based analysis of inpatients treated in 89 German Helios hospitals with laboratory confirmed SARS-CoV-2 infection. ICD-10-GM-code U07.1 (International Statistical Classification of Diseases and Related Health Problems Version 10, German Modification) as main or secondary diagnosis was used to identify COVID-19. Present SARS-CoV-2 infection was linked to the occurrence of SARI, commonly defined by the WHO as an acute respiratory illness with a history of fever, cough, onset within the past 10 days and requiring hospitalization ^17^. SARI cases were identified by ICD-10-codes J09-J22 (main or secondary diagnosis) in accordance with common methods of SARI surveillance ^18^. ICD-10-codes J09-J22 comprise influenza and pneumonia (J09-J18), acute bronchitis (J20), acute bronchiolitis (J21) and unspecified acute lower respiratory tract infection (J22). To evaluate of temporal trends of SARI and COVID-19, we analysed the whole pandemic period from March 2020 to February 2022. Patients with full inpatient treatment during the observational period of nine weeks between December 6, 2021 and February 6, 2022 underwent a detailed analysis. Relevant treatments and patient characteristics were analysed according to the Operation and Procedure Classification System (OPS) and ICD-10-codes based on the Elixhauser comorbidity index ^19^ (Supplemental Table 1). The following outcomes and treatments were analysed: intensive care treatment (OPS-codes 8-980, 8-98f or duration of intensive-care stay > 0), mechanical ventilation (OPS-codes 8-70x, 8-71x or duration of ventilation > 0), in-hospital mortality, length of stay (in days), length of stay at intensive care unit (ICU, in days), duration of mechanical ventilation (hours). For the analysis of in-hospital mortality, we excluded cases with discharge due to hospital transfer or unspecified reasons.

Based on data made available by the RKI ^20^ but assuming a one week delay between infection and consecutive hospital admission, three cohorts and time periods were defined with respect to the dominating virus variant: Delta dominance (2021-12-06 to 2021-12-26), Delta to Omicron transition phase (2021-12-27 to 2022-01-16) and Omicron dominance (2022-01-17 to 2022-02-06).

Inferential statistics were based on generalized linear mixed models (GLMM) specifying hospitals as random factor ^21^. We employed logistic GLMMs function for dichotomous data and linear mixed models (LMM) for continuous data. Effects were estimated with the lme4 package (version 1.1-26) ^22^ in the R environment for statistical computing (version 4.0.2, 64-bit build) ^23^. For all tests we apply a two-tailed 5% error criterion for significance. For the description of the patient characteristics of the cohorts, we employed χ^2^-tests for binary variables and analysis of variance for numeric variables. We report proportions, means, standard deviations, and p-values.

For the comparison of proportions of symptoms as well as selected treatments and outcomes in the different cohorts, we used logistic GLMMs with logit link function. We report proportions, odds ratios together with confidence intervals and p-values. The analysis of the outcome variables length of stay, length of ICU stay, and duration of mechanical ventilation was performed via LMMs. For these analyses, we log-transformed the dependent variables. We report means, standard deviations, medians, interquartile ranges, and p-values. The computation of p-values is based on the Satterthwaite approximation for degrees of freedom.

GLMMs and LMMs were fitted adjusting for age, gender and Elixhauser score. Age and gender are normalized before fitting the models. We report statistics for Elixhauser comorbidity index as well as its items. For the weighted Elixhauser comorbidity index, the Agency for Healthcare Research and Quality (AHRQ) algorithm was applied ^19^.

The analysis was carried out according to the principles outlined in the Declaration of Helsinki. Patient-related data were stored in an anonymized form. The local ethics committee (vote: AZ490/20-ek) and the Helios Kliniken GmbH data protection authority approved data use for this study.

## Results

The central illustration displays trends in COVID-19 diagnoses since the onset of the global pandemic stratified by additionally encoded SARI. During each of the first four pandemic waves, a large proportion of inpatients with COVID-19 also had SARI (hereafter referred to as SARI+). The share of patients with present SARS-CoV-2 infection but without SARI (hereafter referred to as SARI-) mostly followed the course of the pandemic waves but case numbers remained significantly below those with SARI. During the Delta to Omicron transition phase, this relation changed. Case numbers of SARI-exceeded SARI+ for the first time in the pandemic (Figure 1, Delta to Omicron transition phase marked in green).

**Figure 1.**
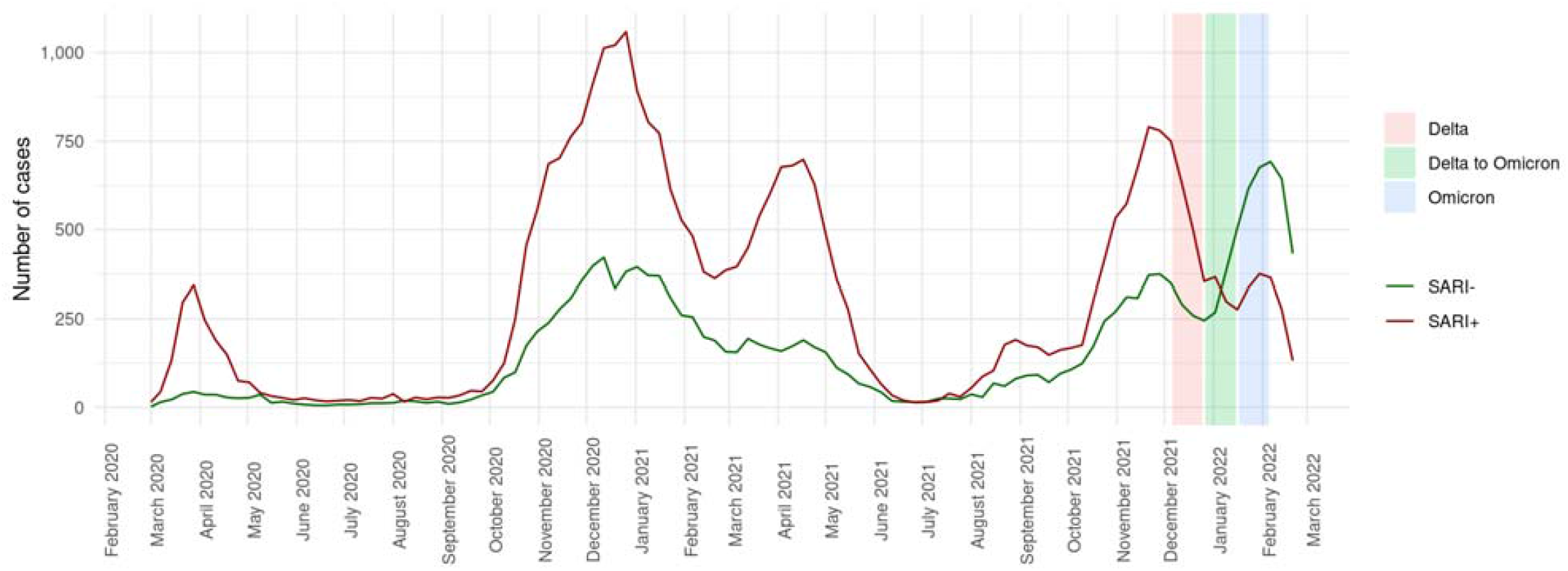
Central illustration: COVID-19 cases since beginning of 2020 stratified by encoded SARI. The coloured bars represent three phases with respect to the dominating SARS-CoV-2 variants. SARI = Severe Acute Respiratory Infection; SARI- = COVID-19 without SARI; SARI+ = COVID-19 with SARI

A total of 4,494 patients with inpatient treatment and laboratory confirmed COVID-19 diagnosis were identified in the period 2021-12-06 to 2022-02-06 (Table 1). Patient characteristics are shown in Supplemental Table 2. Proportion of SARI+ cases decreased from 64.3% in the Delta dominance period to 30.6% in the Omicron dominance period (p<0.01). Mean age [SD] in the total COVID-19 cohort decreased from 61.6 [22.2] to 47.8 [28.1] (p<0.01). This was driven by a significant increase of patients in the age group ≤59 years. Patients in the SARI+ cohort were older as compared to SARI- and mean age also decreased significantly throughout the observational period when the separate cohorts were analysed (Supplemental Table 2). Proportion of male patients decreased significantly for the total cohort (52.5% to 45.3%; p<0.01). Regarding comorbidities, a significant reduction in the mean weighted Elixhauser comorbidity index could be observed for the total cohort with 8.2 [9.4] during Delta and 5.4 [8.7] during Omicron (p<0.01) and the SARI-cohort (6.1 [8.9] to 4.1 [7.8]; p<0.01) but not for SARI+ (9.4 [9.5] to 8.5 [9.8]; p=0.22).

**Table 1:**
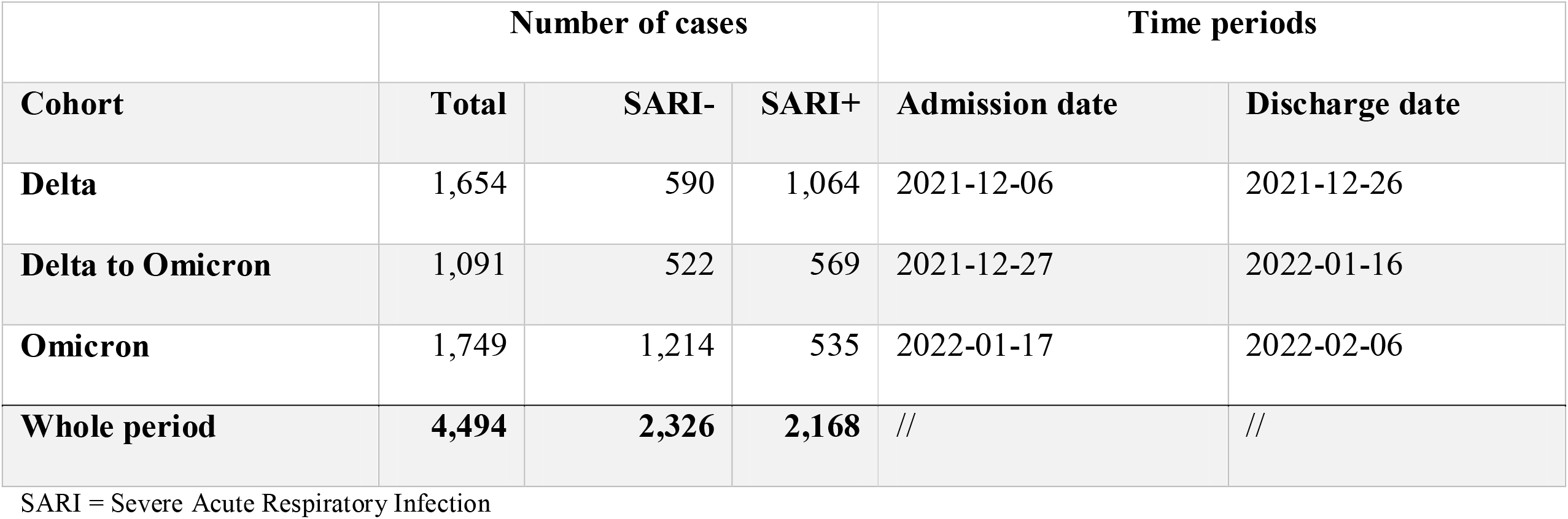
Cohorts and case numbers.

|  | Number of cases |  |  | Time periods |  |
| --- | --- | --- | --- | --- | --- |
| Cohort | Total | SARI- | SARI+ | Admission date | Discharge date |
| Delta | 1,654 | 590 | 1,064 | 2021-12-06 | 2021-12-26 |
| Delta to Omicron | 1,091 | 522 | 569 | 2021-12-27 | 2022-01-16 |
| Omicron | 1,749 | 1,214 | 535 | 2022-01-17 | 2022-02-06 |
| Whole period | 4,494 | 2,326 | 2,168 | // | // |

SARI = Severe Acute Respiratory Infection

For all treatments and outcomes, odds were significantly reduced comparing the Delta and Omicron periods in the total COVID-19 cohort (Table 2). Adjusted odds ratios [95%CI] were 0.64 [0.51-0.80] for intensive care treatment (p<0.001), 0.38 [0.28-0.51] for mechanical ventilation (p<0.001) and 0.42 [0.32-0.56] for in-hospital mortality (p<0.001). Raw in-hospital mortality rates were 16.3% for the Delta period, 12.2% for Delta to Omicron transition phase and 5.5% for the Omicron period. Considering the SARI+ cohort, a similar picture emerged with adjusted odds ratios comparing Delta and Omicron of 0.61 [0.45-0.83] for intensive care treatment (p=0.002), 0.55 [0.39-0.77] for mechanical ventilation (p<0.001) and 0.63 [0.46-0.88] for in-hospital mortality (p=0.006). No significant risk reduction could be observed in the SARI-cohort for intensive care treatment and mechanical ventilation but for in-hospital mortality (p=0.021). Length of stay, length of ICU stay as well as the duration of mechanical ventilation were reduced significantly in the total cohort comparing Delta and Omicron (Table 3), regarding length of stay and duration of mechanical ventilation also in the SARI+ cohort.

**Table 2:** Comparison of outcomes and treatments, adjusted for age, gender and Elixhauser score.

| Cohort | Total |  |  | SARI- |  |  | SARI+ |  |  |
| --- | --- | --- | --- | --- | --- | --- | --- | --- | --- |
|  | Proportion<br>(*n*) | Odds ratio (95%<br>CI) | *P* value | Proportion<br>(*n*) | Odds ratio (95%<br>CI) | *P* value | Proportion<br>(*n*) | Odds ratio (95%<br>CI) | *P* value |
| **Intensive care treatment** |  |  |  |  |  |  |  |  |  |
| **Delta** | 16.6% (275) |  |  | 8.3% (49) |  |  | 21.2% (226) |  |  |
| **Delta to**<br>**Omicron** | 16.2% (177) | 1.11 (0.89-1.38) | 0.372 | 10.9% (57) | 1.56 (1.01-2.40) | 0.046 | 21.1% (120) | 1.08 (0.83-1.40) | 0.585 |
| **Omicron** | 9.5% (167) | 0.64 (0.51-0.80) | < 0.001 | 7.2% (88) | 1.13 (0.75-1.70) | 0.549 | 14.8% (79) | 0.61 (0.45-0.83) | 0.002 |
| **Mechanical ventilation** |  |  |  |  |  |  |  |  |  |
| **Delta** | 12.2% (202) |  |  | 1.0% (6) |  |  | 18.4% (196) |  |  |
| **Delta to**<br>**Omicron** | 10.8% (118) | 1.01 (0.79-1.31) | 0.914 | 0.8% (4) | 0.83 (0.23-3.00) | 0.781 | 20.0% (114) | 1.25 (0.95-1.65) | 0.106 |
| **Omicron** | 4.0% (70) | 0.38 (0.28-0.51) | < 0.001 | 0.9% (11) | 1.15 (0.42-3.18) | 0.789 | 11.0% (59) | 0.55 (0.39-0.77) | < 0.001 |
| **In-hospital mortality\*** |  |  |  |  |  |  |  |  |  |
| **Delta** | 16.3% (254) |  |  | 3.9% (22) |  |  | 23.4% (232) |  |  |
| **Delta to**<br>**Omicron** | 12.2% (126) | 0.90 (0.70-1.17) | 0.436 | 2.8% (14) | 0.95 (0.46-1.94) | 0.888 | 21.1% (112) | 1.02 (0.76-1.36) | 0.911 |
| **Omicron** | 5.5% (94) | 0.42 (0.32-0.56) | < 0.001 | 1.1% (13) | 0.43 (0.21-0.88) | 0.021 | 15.7% (81) | 0.63 (0.46-0.88) | 0.006 |

\*based on 4,286 cases (95.4%). We excluded cases with discharge due to hospital transfer or unspecified reason. CI = Confidence interval; SARI = Severe Acute Respiratory Infection

**Table 3:** Length of stay, adjusted for age, gender and Elixhauser score.

| Cohort | Total |  |  |  | SARI- |  |  |  | SARI+ |  |  |  |
| --- | --- | --- | --- | --- | --- | --- | --- | --- | --- | --- | --- | --- |
|  | Mean (SD) | Median (IQR) | Coefficient (95% CI) | P value | Mean (SD) | Median (IQR) | Coefficient (95% CI) | P value | Mean (SD) | Median (IQR) | Coefficient (95% CI) | P value |
| **Length of stay** |  |  |  |  |  |  |  |  |  |  |  |  |
| **Delta** | 5.5 ±3.8 | 4.0 [3, 7] |  |  | 4.1 ±3.3 | 3.0 [2, 5] |  |  | 6.3 ±3.8 | 6.0 [3, 8] |  |  |
| **Delta to Omicron** | 4.9 ±3.8 | 4.0 [2, 7] | -0.06 (-0.10 to -0.02) | < 0.01 | 3.6 ±3.1 | 3.0 [2, 5] | -0.03 (-0.09 to 0.03) | 0.29 | 6.1 ±4.1 | 5.0 [3, 8] | -0.03 (-0.08 to 0.03) | 0.35 |
| **Omicron** | 3.9 ±3.1 | 3.0 [2, 5] | -0.16 (-0.20 to -0.12) | < 0.01 | 3.4 ±2.8 | 2.0 [2, 4] | -0.04 (-0.09 to 0.01) | 0.16 | 5.1 ±3.6 | 4.0 [2, 7] | -0.16 (-0.21 to -0.10) | < 0.01 |
| **Length of stay at ICU\*** |  |  |  |  |  |  |  |  |  |  |  |  |
| **Delta** | 4.5 ±3.6 | 3.0 [2, 6] |  |  | 2.7 ±2.3 | 2.0 [1, 3] |  |  | 4.9 ±3.8 | 4.0 [2, 7] |  |  |
| **Delta to Omicron** | 4.2 ±3.7 | 3.0 [1, 6] | -0.11 (-0.27 to 0.05) | 0.16 | 1.8 ±1.3 | 1.0 [1, 2] | -0.32 (-0.54 to -0.09) | < 0.01 | 5.3 ±4.0 | 4.0 [2, 7] | 0.09 (-0.09 to 0.27) | 0.34 |
| **Omicron** | 3.2 ±3.2 | 2.0 [1, 4] | -0.39 (-0.55 to -0.23) | < 0.01 | 1.9 ±1.5 | 1.0 [1, 2] | -0.26 (-0.47 to -0.05) | 0.02 | 4.5 ±3.9 | 3.0 [2, 6] | -0.15 (-0.36 to 0.06) | 0.17 |
| **Duration of mechanical ventilation†** |  |  |  |  |  |  |  |  |  |  |  |  |
| **Delta** | 104.7 ±87.9 | 83.0 [37, 149] |  |  | 67.7 ±63.5 | 70.0 [36, 100] |  |  | 105.4 ±88.3 | 83.5 [37, 150] |  |  |
| **Delta to Omicron** | 95.1 ±95.5 | 58.0 [22, 154] | -0.36 (-0.71 to -0.01) | 0.05 | 23.7 ±18.6 | 26.0 [15, 34] | -1.76 (-3.93 to 0.41) | 0.16 | 97.5 ±96.1 | 61.0 [22, 154] | -0.34 (-0.70 to 0.02) | 0.06 |
| **Omicron** | 78.0 ±88.3 | 41.0 [12, 135] | -0.63 (-1.08 to -0.19) | < 0.01 | 42.9 ±30.8 | 36.0 [26, 52] | -0.13 (-1.90 to 1.64) | 0.89 | 83.8 ±93.5 | 41.5 [10, 154] | -0.63 (-1.11 to -0.16) | < 0.01 |

\* Based on 1,234 cases (13.7%). We excluded cases with length of stay at ICU = 0; † Based on 598 cases (6.7%). We excluded cases with duration of ventilation = 0; CI = Confidence interval; SARI = Severe Acute Respiratory Infection

## Discussion

We present an analysis of 4,494 inpatients treated in 89 German hospitals with laboratory confirmed SARS-CoV-2 infection during the transition period of dominating virus variants Delta and Omicron. To the best of our knowledge, this is the first study comparing patient outcomes of COVID-19 during the fourth and fifth pandemic wave in Germany with respect to the dominating VoC.

Our analysis yielded the following findings: Occurrence of SARI is common among COVID-19 patients which was most obvious during the first four pandemic waves. During the Delta to Omicron transition phase, case numbers of SARI-patients exceeded those of SARI+. We noticed a significant change in patient characteristics. In the phase of Omicron dominance, patients were younger, more likely to be female and characterized by a lower comorbidity burden. Patients infected during the Omicron dominance period compared to Delta were less likely to receive intensive care treatment, mechanical ventilation and were at significantly lower mortality risk in the total cohort as well as the SARI+ cohort. This leads to the assumption that disease severity in Omicron might be lower. Concomitant occurrence of SARI in hospitalized patients seems to correlate with the severity of SARS-CoV-2 infections. SARI surveillance could therefore play a crucial role in pandemic monitoring. However, future studies are needed to confirm this observation.

Our results are in line with the findings by Nyberg *et al*. ^15^ and other previously mentioned studies ^12-14^ as to the dominant SARS-CoV-2 variant Omicron appears to cause milder courses of infection. Importantly, Nyberg *et al*. performed stratifications by vaccination status and history of past infection which confirmed the reduction of disease severity even in unvaccinated subjects. As administrative data does not contain information about vaccination status, a similar stratification could not be executed within our analysis. Therefore, potential influence of an underlying population immunity which prevents severe courses of infection cannot be ruled out. The current pandemic situation in Hong Kong shows that Omicron can indeed lead to high case fatality rates in a rather non-immunized population due to low vaccination rates and low background immunity because of the so-called “Zero-COVID strategy” ^24^. The effect of booster vaccinations, which offer 70% protection against hospitalisation and death as was reported Nyberg *et al*.^15^, must also be highlighted as a large proportion of booster doses were administered during the Omicron period ^14^. However, our observation that patients in the SARI+ group had a comparable comorbidity burden during Omicron and Delta phases (as indicated by Elixhauser comorbidity index values) but still were at lower risk for adverse outcomes strengthens the assumption of a reduced disease severity which is also backed by a recent experimental study available as a preprint ^25^.

According to our data, Omicron affects a younger population, more frequently females and patients with lower comorbidity burden. A similar shift in patient characteristics was observed in previous studies investigating the clinical differences of Omicron and Delta – for example in a recent study by Bouzid *et al*. who analysed 1,716 patients presenting to 13 emergency departments in France ^26^. In this analysis, prevalence of comorbidities (except obesity) did not differ between the groups, however, only a few distinct comorbidities were surveyed. With respect to outcomes, the findings were comparable to our analysis with reduced risk among Omicron patients for ICU treatment, mechanical ventilation and in-hospital death ^26^. A possible explanation for the shift in demographic parameters, especially age, might be the proportion of booster vaccinations among people aged 60+ which is significantly higher in comparison to people in the age group 18-59 according to RKI data ^27^.

Leveraging existing national and international surveillance systems to monitor pandemic trends is of crucial importance as was also stated by the WHO and ECDC ^28-31^. In Germany, an ICD-code-based SARI surveillance system (ICOSARI) closely linked to the RKI was introduced in 2017 ^18^. In a recent preprint article, Tolksdorf *et al*. analysed hospitalization numbers and ICU treatments derived from ICOSARI in the first three pandemic waves, compared those with the German national notification system for SARS-CoV-2 infections and found a more accurate estimation of hospitalizations and ICU treatments utilizing ICOSARI data ^30^. Monitoring trends of SARI-COVID cases has yet become an integral part of pandemic surveillance in Germany ^16^. Our study further highlights, that this easily applicable system fully based on administrative data could play a central role in assessing disease severity.

### Limitations

We acknowledge several limitations in connection with this study. First, some limitations must be attributed to administrative data in general as data quality depends on correct coding and data is stored for remuneration reasons. However, SARI surveillance in Germany utilizing claims data is a robust and valid approach as was mentioned previously ^18^. Second, laboratory-confirmed COVID-19 cases were identified through ICD-10-codes and no data for viral genome sequencing was available which could have potentially biased the results. Thus, identification of Omicron and Delta cases were based on epidemiological data provided by the RKI ^16 32^. In addition, differentiation between BA.1 and BA.2 sub-lineages was not possible. Third, as outlined above, the effect of person-level vaccination status could not be assessed in this study as respective data was not available. Fourth, identification of COVID-19 cases is dependent on correct testing and cases might stay undetected. However, in Helios hospitals, reverse transcriptase polymerase chain reaction (RT-PCR) testing is mandatory for inpatients in addition to rapid antigen tests. Fifth, nosocomial COVID-19 cases were not excluded in the present analysis. The increased transmissibility of Omicron could have led to more incident infections with unknown impact on disease severity assessment ^14^. Sixth, this analysis included all patients hospitalized within the observational period with a documented COVID-19 diagnosis. The reason for admission was therefore not considered which may have led to possible selection bias as, due to postponement of elective procedures, relatively more patients with severe diseases that require urgent treatment entered the healthcare during the fourth and fifth pandemic wave.

## Conclusion

During the Omicron dominance period, numbers of SARI in hospitalized patients with COVID-19 were lower relative to Non-SARI cases. Analysis of timely trends revealed that this represents a unique finding during the pandemic’s course. Adjusted for age, gender and comorbidity burden, the risk for adverse hospital outcomes (ICU treatment, mechanical ventilation, and in-hospital mortality) was substantially decreased during Omicron dominance as compared to Delta. Patients affected by Omicron were younger, more likely to be female and the comorbidity burden was lower. Continuous ICD-code based SARI monitoring could contribute decisively to COVID-19 surveillance and evaluation of disease severity especially in the light of possible future virus variants.

## Data Availability

The data that support the findings of this study are not publicly available as they contain information that could compromise the privacy of research participants but are available from the corresponding author, Mr. Johannes Leiner upon reasonable request.

## Abbreviations

CI: Confidence interval
COVID-19: Coronavirus disease 2019
ECDC: European Centre for Disease Prevention and Control
GLMM: Generalized linear mixed models
ICD-10-GM: International Statistical Classification of Diseases and Related Health Problems Version 10, German Modification
ICU: Intensive care unit
LMM: Linear mixed models
OPS: Operation and Procedure Classification System
RKI: Robert-Koch-Institute
SARI: Severe acute respiratory infection
SARI+: COVID-19 with SARI
SARI-: COVID-19 without SARI
SARS-CoV-2: Severe acute respiratory syndrome coronavirus 2
VoC: Variant of Concern
WHO: World Health Organization

## Guarantor statement

Mr. Johannes Leiner (JL) takes full responsibility for the content of the manuscript, including the data and analysis.

## Author contributions

VP, SH, SK, ES, RM, GH, RK, AB contributed substantially to the study design, data analysis and interpretation and revision of the manuscript and gave final approval for publishing.

## Statement of Ethics

The analysis was carried out according to the principles outlined in the Declaration of Helsinki. Patient-related data were stored in a pseudonymized form. The local ethics committee (vote: AZ490/20-ek) and the Helios Kliniken GmbH data protection authority approved data use for this study.

## Supplementary Material

Supplemental Table 1: ICD-10-GM-codes used to calculate Elixhauser comorbidity score (according to Moore et al., 2017)

Supplemental Table 2: Baseline characteristics

