## Supplementary material for "Characteristics and outcomes of COVID-19 patients during B.1.1.529 (Omicron) dominance compared to B.1.617.2 (Delta) in 89 German hospitals"

Supplemental Table 1: ICD-10-GM-codes used to calculate Elixhauser comorbidity index (according to Moore et al., 2017)

| Elixhauser comorbidity | Weight | ICD-10-GM-code |
| --- | --- | --- |
| AIDS / HIV | 0 | B20, B21, B22, B23, B24 |
| Alcohol Abuse | -1 | F10, E52, G62.1, I42.6, K29.2, K70.0, K70.3, K70.9, T51, Z50.2, Z71.4, Z72.1 |
| Blood Loss Anemia | -3 | D50.0 |
| Cardiac Arrhythmia | 0 | I44.1, I44.2, I44.3, I45.6, I47, I48, I49, R00.0, R00.1, R00.8, T82.1, Z45.00, Z45.01, Z95.0 |
| Chronic Pulmonary Disease | 3 | I27.8, I27.9, J40, J41, J42, J43, J44, J45, J46, J47, J60, J61, J62, J63, J64, J65, J66, J67, J68.4, J70.1, J70.3 |
| Chronic Renal Failure | 6 | I12.0, I31.1, N18, N19, N25.0, Z49.0, Z49.1, Z49.2, Z94.0, Z99.2 |
| Coagulopathy | 11 | D65, D66, D67, D68, D69.1, D69.3, D69.4, D69.5, D69.6 |
| Congestive Heart Failure | 9 | I09.0, I11.0, I13.0, I13.2, I25.5, I42.0, I42.1, I42.2, I42.5, I42.6, I42.7, I42.8, I42.9, I43, I50 |
| Deficiency Anemia | -2 | D50.8, D50.9, D51, D52, D53 |
| Depression | -5 | F20.4, F31.3 - F31.5, F32, F33, F34.1, F41.2, F43.2 |
| Diabetes Mellitus, Uncomplicated | 0 | E10.0, E10.1, E10.9, E11.0, E11.1, E11.9, E12.0, E12.1, E12.9, E13.0, E13.1, E13.9, E14.0, E14.1, E14.9 (excluding E10.2, E10.3, E10.4, E10.5, E10.6, E10.7, E10.8, E11.2, E11.3, E11.4, E11.5, E11.6, E11.7, E11.8, E12.2, E12.3, E12.4, E12.5, E12.6, E12.7, E12.8, E13.2, E13.3, E13.4, E13.5, E13.6, E13.7, E13.8, E14.2, E14.3, E14.4, E14.5, E14.6, E14.7, E14.8) |
| Diabetes Mellitus, Complicated | -3 | E10.2, E10.3, E10.4, E10.5, E10.6, E10.7, E10.8, E11.2, E11.3, E11.4, E11.5, E11.6, E11.7, E11.8, E12.2, E12.3, E12.4, E12.5, E12.6, E12.7, E12.8, E13.2, E13.3, E13.4, E13.5, E13.6, E13.7, E13.8, E14.2, E14.3, E14.4, E14.5, E14.6, E14.7, E14.8 |
| Drug Abuse | -7 | F11, F12, F13, F14, F15, F16, F18, F19, Z71.5, Z72.2 |
| Fluid And Electrolyte Disorders | 11 | E22.2, E86, E87 |
| Hypertension (combined uncomplicated and complicated) | -1 | I10, I11, I12, I13, I15 |
| Hypothyroidism | 0 | E00, E01, E02, E03, E89.0 |
| Liver Disease | 4 | B18, I85, I86.4, I98.2, K70, K71.1, K71.3, K71.4, K71.5, K71.7, K72, K73, K74, K76.0, K76.2, K76.9, Z94.4 |
| Lymphoma | 6 | C81, C82, C83, C84, C85, C88, C96, C90.0, C90.2 |
| Metastatic Cancer | 14 | C77, C78, C79, C80 |
| Neurological Disorders, other | 5 | G10, G11, G12, G13. G20, G21, G22, G25.4, G25.5, G31.2, G31.8, G31.9, G32, G35, G36, G37, G40, G41, G93.1, G93.4, R47.0, R56 |
| Obesity | -5 | E66 |
| Paralysis | 5 | G04.1, G11.4, G80.1, G80.2, G81, G82, G83.0, G83.1, G83.2, G83.3, G83.4, G83.9 |
| Peptic Ulcer Disease, Excluding Bleeding | 0 | K25.7, K25.9, K26.7, K26.9, K27.7, K27.9, K28.7, K28.9 |
| Peripheral Vascular Disorders | 3 | I70, I71, I73.1, I73.8, I73.9, I77.1, I79.0, I79.2, Z95.81, Z95.88, Z95.9 |
| Psychoses | -5 | F20, F22, F23, F24, F25, F28, F29, F30.2, F31.2, F31.5 |
| Pulmonary Circulation Disorders | 6 | I26, I27, I28.0, I28.8, I28.9 |
| Rheumatoid Arthritis / Collagen Vascular Diseases | 0 | L94.0, L94.1, L94.3, M05, M06, M08, M12.0, M12.3, M30, M31.0, M31.1, M31.2, M31.3, M32, M33, M34, M35, M45, M46.1, M46.8, M46.9 |
| Solid Tumor Without Metastases | 7 | C00, C01, C02, C03, C04, C05, C06, C07, C08, C09, C10, C11, C12, C13, C14, C15, C16, C17, C18, C19, C20, C21, C22, C23, C24, C25, C26, C30, C31, C32, C33, C34, C37, C38, C39, C40, C41, C43, C45, C46, C47, C48, C49, C50, C51, C52, C53, C54, C55, C56, C57, C58, C60, C61, C62, C63, C64, C65, C66, C67, C68, C69, C70, C71, C72, C73, C74, C75, C76, C97 |
| Valvular Heart Disease | 0 | I05, I06, I07, I08, I09.1, I34, I35, I36, I37, I38, I39, Q23.0, Q23.1, Q23.2, Q23.3, Z95.2, Z95.3, Z95.4 |
| Weight Loss | 9 | E40, E41, E42, E43, E44, E45, E46, R63.4, R64 |

ICD-10-GM = German Modification of the International Statistical Classification of Diseases and Related Health Problems Version 10

Supplemental Table 2: Baseline characteristics

|  | Total | | | | SARI- | | | | SARI+ | | | |
| --- | --- | --- | --- | --- | --- | --- | --- | --- | --- | --- | --- | --- |
|  | **Proportion (*n*)** | | |  | **Proportion (*n*)** | | |  | **Proportion (*n*)** | | |  |
| Group | **Delta** | **Delta to Omicron** | **Omicron** | ***P* value** | **Delta** | **Delta to Omicron** | **Omicron** | ***P* value** | **Delta** | **Delta to Omicron** | **Omicron** | ***P* value** |
| Age | | | | | | | | | | | | |
| *Mean (SD)* | 61.6 ±22.2 | 54.9 ±25.4 | 47.8 ±28.1 | < 0.01 | 53.9 ±26.9 | 46.3 ±27.0 | 43.2 ±27.5 | < 0.01 | 65.8 ±17.7 | 62.9 ±20.9 | 58.3 ±26.6 | < 0.01 |
| ≤ 59 years | 40.0% (662) | 49.2% (537) | 60.5% (1,058) | < 0.01 | 51.2% (302) | 62.3% (325) | 68.2% (828) | < 0.01 | 33.8% (360) | 37.3% (212) | 43.0% (230) | < 0.01 |
| 60−69 years | 17.3% (286) | 16.1% (176) | 10.0% (175) | < 0.01 | 13.4% (79) | 12.8% (67) | 8.4% (102) | < 0.01 | 19.5% (207) | 19.2% (109) | 13.6% (73) | 0.01 |
| 70−79 years | 18.3% (302) | 14.0% (153) | 11.4% (200) | < 0.01 | 13.6% (80) | 10.2% (53) | 9.2% (112) | 0.02 | 20.9% (222) | 17.6% (100) | 16.4% (88) | 0.07 |
| ≥ 80 years | 24.4% (404) | 20.6% (225) | 18.1% (316) | < 0.01 | 21.9% (129) | 14.8% (77) | 14.2% (172) | < 0.01 | 25.8% (275) | 26.0% (148) | 26.9% (144) | 0.90 |
| Sex | | | | | | | | | | | | |
| Male | 52.5% (869) | 47.0% (513) | 45.3% (793) |  | 44.1% (260) | 41.4% (216) | 41.4% (503) |  | 57.2% (609) | 52.2% (297) | 54.2% (290) |  |
| Female | 47.5% (785) | 53.0% (578) | 54.7% (956) | < 0.01 | 55.9% (330) | 58.6% (306) | 58.6% (711) | 0.53 | 42.8% (455) | 47.8% (272) | 45.8% (245) | 0.13 |
| SARI | | | | | | | | | | | | |
| No SARI | 35.7% (590) | 47.8% (522) | 69.4% (1,214) |  | 100.0% (590) | 100.0% (522) | 100.0% (1,214) |  | 0.0% (0) | 0.0% (0) | 0.0% (0) |  |
| SARI | 64.3% (1,064) | 52.2% (569) | 30.6% (535) | < 0.01 | 0.0% (0) | 0.0% (0) | 0.0% (0) |  | 100.0% (1,064) | 100.0% (569) | 100.0% (535) |  |
| Elixhauser comorbidity index | | | | | | | | | | | | |
| *Mean (SD)* | 8.2 ± 9.4 | 6.9 ± 9.3 | 5.4 ± 8.7 | < 0.01 | 6.1 ± 8.9 | 4.6 ± 9.0 | 4.1 ± 7.8 | < 0.01 | 9.4 ± 9.5 | 9.1 ± 9.2 | 8.5 ± 9.8 | 0.22 |
| < 0 | 13.2% (219) | 15.7% (171) | 13.3% (233) | 0.14 | 15.8% (93) | 19.3% (101) | 14.1% (171) | 0.02 | 11.8% (126) | 12.3% (70) | 11.6% (62) | 0.93 |
| 0 | 22.5% (372) | 31.1% (339) | 41.3% (723) | < 0.01 | 31.7% (187) | 43.7% (228) | 48.0% (583) | < 0.01 | 17.4% (185) | 19.5% (111) | 26.2% (140) | < 0.01 |
| 1-4 | 6.3% (104) | 3.6% (39) | 5.1% (90) | < 0.01 | 6.8% (40) | 4.0% (21) | 4.6% (56) | 0.07 | 6.0% (64) | 3.2% (18) | 6.4% (34) | 0.03 |
| ≥ 5 | 58.0% (959) | 49.7% (542) | 40.2% (703) | < 0.01 | 45.8% (270) | 33.0% (172) | 33.3% (404) | < 0.01 | 64.8% (689) | 65.0% (370) | 55.9% (299) | < 0.01 |
|  | **Total** | | | | **SARI-** | | | | **SARI+** | | | |
| Group | **Delta** | **Delta to Omicron** | **Omicron** | ***P* value** | **Delta** | **Delta to Omicron** | **Omicron** | ***P* value** | **Delta** | **Delta to Omicron** | **Omicron** | ***P* value** |
| Congestive heart failure | | | | | | | | | | | | |
| no | 82.7% (1,368) | 87.5% (955) | 89.4% (1,564) |  | 88.0% (519) | 90.0% (470) | 92.1% (1,118) |  | 79.8% (849) | 85.2% (485) | 83.4% (446) |  |
| yes | 17.3% (286) | 12.5% (136) | 10.6% (185) | < 0.01 | 12.0% (71) | 10.0% (52) | 7.9% (96) | 0.02 | 20.2% (215) | 14.8% (84) | 16.6% (89) | 0.02 |
| Cardiac arrhythmias | | | | | | | | | | | | |
| no | 80.4% (1,330) | 83.0% (905) | 87.2% (1,526) |  | 82.5% (487) | 87.7% (458) | 89.6% (1,088) |  | 79.2% (843) | 78.6% (447) | 81.9% (438) |  |
| yes | 19.6% (324) | 17.0% (186) | 12.8% (223) | < 0.01 | 17.5% (103) | 12.3% (64) | 10.4% (126) | < 0.01 | 20.8% (221) | 21.4% (122) | 18.1% (97) | 0.34 |
| Valvular disease | | | | | | | | | | | | |
| no | 94.9% (1,570) | 96.5% (1,053) | 96.8% (1,693) |  | 95.9% (566) | 96.0% (501) | 96.9% (1,176) |  | 94.4% (1,004) | 97.0% (552) | 96.6% (517) |  |
| yes | 5.1% (84) | 3.5% (38) | 3.2% (56) | 0.01 | 4.1% (24) | 4.0% (21) | 3.1% (38) | 0.49 | 5.6% (60) | 3.0% (17) | 3.4% (18) | 0.02 |
| Pulmonary circulation disorders | | | | | | | | | | | | |
| no | 94.7% (1,566) | 95.6% (1,043) | 97.9% (1,713) |  | 97.1% (573) | 98.5% (514) | 98.5% (1,196) |  | 93.3% (993) | 93.0% (529) | 96.6% (517) |  |
| yes | 5.3% (88) | 4.4% (48) | 2.1% (36) | < 0.01 | 2.9% (17) | 1.5% (8) | 1.5% (18) | 0.10 | 6.7% (71) | 7.0% (40) | 3.4% (18) | 0.01 |
| Peripheral vascular disorders | | | | | | | | | | | | |
| no | 94.9% (1,569) | 95.2% (1,039) | 96.3% (1,684) |  | 93.9% (554) | 96.2% (502) | 96.6% (1,173) |  | 95.4% (1,015) | 94.4% (537) | 95.5% (511) |  |
| yes | 5.1% (85) | 4.8% (52) | 3.7% (65) | 0.12 | 6.1% (36) | 3.8% (20) | 3.4% (41) | 0.02 | 4.6% (49) | 5.6% (32) | 4.5% (24) | 0.60 |
| Hypertension, uncomplicated | | | | | | | | | | | | |
| no | 63.3% (1,047) | 68.2% (744) | 74.2% (1,298) |  | 66.6% (393) | 74.1% (387) | 78.2% (949) |  | 61.5% (654) | 62.7% (357) | 65.2% (349) |  |
| yes | 36.7% (607) | 31.8% (347) | 25.8% (451) | < 0.01 | 33.4% (197) | 25.9% (135) | 21.8% (265) | < 0.01 | 38.5% (410) | 37.3% (212) | 34.8% (186) | 0.34 |
| Hypertension, complicated | | | | | | | | | | | | |
| no | 89.5% (1,480) | 93.4% (1,019) | 93.9% (1,642) |  | 91.0% (537) | 95.2% (497) | 95.3% (1,157) |  | 88.6% (943) | 91.7% (522) | 90.7% (485) |  |
| yes | 10.5% (174) | 6.6% (72) | 6.1% (107) | < 0.01 | 9.0% (53) | 4.8% (25) | 4.7% (57) | < 0.01 | 11.4% (121) | 8.3% (47) | 9.3% (50) | 0.11 |
| Paralysis | | | | | | | | | | | | |
| no | 96.9% (1,603) | 97.1% (1,059) | 97.7% (1,709) |  | 96.4% (569) | 96.6% (504) | 98.0% (1,190) |  | 97.2% (1,034) | 97.5% (555) | 97.0% (519) |  |
| yes | 3.1% (51) | 2.9% (32) | 2.3% (40) | 0.33 | 3.6% (21) | 3.4% (18) | 2.0% (24) | 0.08 | 2.8% (30) | 2.5% (14) | 3.0% (16) | 0.86 |
|  | **Total** | | | | **SARI-** | | | | **SARI+** | | | |
| Group | **Delta** | **Delta to Omicron** | **Omicron** | ***P* value** | **Delta** | **Delta to Omicron** | **Omicron** | ***P* value** | **Delta** | **Delta to Omicron** | **Omicron** | ***P* value** |
| Other neurological disorders | | | | | | | | | | | | |
| no | 93.5% (1,547) | 94.0% (1,025) | 93.7% (1,638) |  | 93.4% (551) | 94.4% (493) | 93.7% (1,138) |  | 93.6% (996) | 93.5% (532) | 93.5% (500) |  |
| yes | 6.5% (107) | 6.0% (66) | 6.3% (111) | 0.91 | 6.6% (39) | 5.6% (29) | 6.3% (76) | 0.76 | 6.4% (68) | 6.5% (37) | 6.5% (35) | 0.99 |
| Chronic pulmonary disease | | | | | | | | | | | | |
| no | 89.9% (1,487) | 93.8% (1,023) | 92.5% (1,617) |  | 92.7% (547) | 96.0% (501) | 94.0% (1,141) |  | 88.3% (940) | 91.7% (522) | 89.0% (476) |  |
| yes | 10.1% (167) | 6.2% (68) | 7.5% (132) | < 0.01 | 7.3% (43) | 4.0% (21) | 6.0% (73) | 0.07 | 11.7% (124) | 8.3% (47) | 11.0% (59) | 0.10 |
| Diabetes, uncomplicated | | | | | | | | | | | | |
| no | 84.5% (1,398) | 87.5% (955) | 90.2% (1,577) |  | 87.3% (515) | 92.0% (480) | 91.9% (1,116) |  | 83.0% (883) | 83.5% (475) | 86.2% (461) |  |
| yes | 15.5% (256) | 12.5% (136) | 9.8% (172) | < 0.01 | 12.7% (75) | 8.0% (42) | 8.1% (98) | < 0.01 | 17.0% (181) | 16.5% (94) | 13.8% (74) | 0.25 |
| Diabetes, complicated | | | | | | | | | | | | |
| no | 92.1% (1,524) | 93.6% (1,021) | 95.3% (1,667) |  | 94.1% (555) | 95.4% (498) | 97.1% (1,179) |  | 91.1% (969) | 91.9% (523) | 91.2% (488) |  |
| yes | 7.9% (130) | 6.4% (70) | 4.7% (82) | < 0.01 | 5.9% (35) | 4.6% (24) | 2.9% (35) | < 0.01 | 8.9% (95) | 8.1% (46) | 8.8% (47) | 0.84 |
| Hypothyroidism | | | | | | | | | | | | |
| no | 89.8% (1,486) | 91.2% (995) | 92.3% (1,615) |  | 90.0% (531) | 91.8% (479) | 93.2% (1,132) |  | 89.8% (955) | 90.7% (516) | 90.3% (483) |  |
| yes | 10.2% (168) | 8.8% (96) | 7.7% (134) | 0.04 | 10.0% (59) | 8.2% (43) | 6.8% (82) | 0.05 | 10.2% (109) | 9.3% (53) | 9.7% (52) | 0.83 |
| Renal failure | | | | | | | | | | | | |
| no | 78.1% (1,292) | 82.6% (901) | 84.9% (1,485) |  | 83.7% (494) | 88.1% (460) | 89.1% (1,082) |  | 75.0% (798) | 77.5% (441) | 75.3% (403) |  |
| yes | 21.9% (362) | 17.4% (190) | 15.1% (264) | < 0.01 | 16.3% (96) | 11.9% (62) | 10.9% (132) | < 0.01 | 25.0% (266) | 22.5% (128) | 24.7% (132) | 0.51 |
| Liver disease | | | | | | | | | | | | |
| no | 96.9% (1,602) | 97.0% (1,058) | 98.1% (1,715) |  | 97.1% (573) | 96.6% (504) | 98.6% (1,197) |  | 96.7% (1,029) | 97.4% (554) | 96.8% (518) |  |
| yes | 3.1% (52) | 3.0% (33) | 1.9% (34) | 0.06 | 2.9% (17) | 3.4% (18) | 1.4% (17) | 0.01 | 3.3% (35) | 2.6% (15) | 3.2% (17) | 0.76 |
| Peptic ulcer disease excluding bleeding | | | | | | | | | | | | |
| no | 99.9% (1,653) | 99.8% (1,089) | 99.9% (1,748) |  | 99.8% (589) | 99.8% (521) | 99.9% (1,213) |  | 100.0% (1,064) | 99.8% (568) | 100.0% (535) |  |
| yes | 0.1% (1) | 0.2% (2) | 0.1% (1) | 0.49 | 0.2% (1) | 0.2% (1) | 0.1% (1) | 0.80 | 0.0% (0) | 0.2% (1) | 0.0% (0) | 0.25 |
|  | **Total** | | | | **SARI-** | | | | **SARI+** | | | |
| Group | **Delta** | **Delta to Omicron** | **Omicron** | ***P* value** | **Delta** | **Delta to Omicron** | **Omicron** | ***P* value** | **Delta** | **Delta to Omicron** | **Omicron** | ***P* value** |
| AIDS/HIV | | | | | | | | | | | | |
| no | 100.0% (1,654) | 100.0% (1,091) | 100.0% (1,749) |  | 100.0% (590) | 100.0% (522) | 100.0% (1,214) |  | 100.0% (1,064) | 100.0% (569) | 100.0% (535) |  |
| yes | 0.0% (0) | 0.0% (0) | 0.0% (0) |  | 0.0% (0) | 0.0% (0) | 0.0% (0) |  | 0.0% (0) | 0.0% (0) | 0.0% (0) |  |
| Lymphoma | | | | | | | | | | | | |
| no | 98.9% (1,636) | 98.7% (1,077) | 99.2% (1,735) |  | 98.6% (582) | 98.7% (515) | 99.3% (1,205) |  | 99.1% (1,054) | 98.8% (562) | 99.1% (530) |  |
| yes | 1.1% (18) | 1.3% (14) | 0.8% (14) | 0.44 | 1.4% (8) | 1.3% (7) | 0.7% (9) | 0.35 | 0.9% (10) | 1.2% (7) | 0.9% (5) | 0.84 |
| Metastatic cancer | | | | | | | | | | | | |
| no | 98.7% (1,632) | 98.6% (1,076) | 98.5% (1,723) |  | 97.5% (575) | 97.5% (509) | 98.5% (1,196) |  | 99.3% (1,057) | 99.6% (567) | 98.5% (527) |  |
| yes | 1.3% (22) | 1.4% (15) | 1.5% (26) | 0.92 | 2.5% (15) | 2.5% (13) | 1.5% (18) | 0.20 | 0.7% (7) | 0.4% (2) | 1.5% (8) | 0.08 |
| Solid tumour without metastasis | | | | | | | | | | | | |
| no | 95.9% (1,587) | 96.6% (1,054) | 96.0% (1,679) |  | 93.9% (554) | 95.0% (496) | 96.0% (1,166) |  | 97.1% (1,033) | 98.1% (558) | 95.9% (513) |  |
| yes | 4.1% (67) | 3.4% (37) | 4.0% (70) | 0.64 | 6.1% (36) | 5.0% (26) | 4.0% (48) | 0.12 | 2.9% (31) | 1.9% (11) | 4.1% (22) | 0.10 |
| Rheumatoid artritis/collaged vascular disease | | | | | | | | | | | | |
| no | 98.2% (1,624) | 98.7% (1,077) | 98.5% (1,723) |  | 98.1% (579) | 98.5% (514) | 98.7% (1,198) |  | 98.2% (1,045) | 98.9% (563) | 98.1% (525) |  |
| yes | 1.8% (30) | 1.3% (14) | 1.5% (26) | 0.52 | 1.9% (11) | 1.5% (8) | 1.3% (16) | 0.67 | 1.8% (19) | 1.1% (6) | 1.9% (10) | 0.46 |
| Coagulopathy | | | | | | | | | | | | |
| no | 96.4% (1,595) | 97.0% (1,058) | 97.9% (1,713) |  | 97.8% (577) | 97.3% (508) | 97.7% (1,186) |  | 95.7% (1,018) | 96.7% (550) | 98.5% (527) |  |
| yes | 3.6% (59) | 3.0% (33) | 2.1% (36) | 0.03 | 2.2% (13) | 2.7% (14) | 2.3% (28) | 0.86 | 4.3% (46) | 3.3% (19) | 1.5% (8) | 0.01 |
| Obesity | | | | | | | | | | | | |
| no | 88.6% (1,465) | 90.8% (991) | 92.7% (1,622) |  | 93.9% (554) | 92.1% (481) | 93.7% (1,138) |  | 85.6% (911) | 89.6% (510) | 90.5% (484) |  |
| yes | 11.4% (189) | 9.2% (100) | 7.3% (127) | < 0.01 | 6.1% (36) | 7.9% (41) | 6.3% (76) | 0.41 | 14.4% (153) | 10.4% (59) | 9.5% (51) | < 0.01 |
| Weight loss | | | | | | | | | | | | |
| no | 95.6% (1,582) | 94.1% (1,027) | 97.0% (1,696) |  | 95.6% (564) | 96.4% (503) | 98.2% (1,192) |  | 95.7% (1,018) | 92.1% (524) | 94.2% (504) |  |
| yes | 4.4% (72) | 5.9% (64) | 3.0% (53) | < 0.01 | 4.4% (26) | 3.6% (19) | 1.8% (22) | < 0.01 | 4.3% (46) | 7.9% (45) | 5.8% (31) | 0.01 |
|  | **Total** | | | | **SARI-** | | | | **SARI+** | | | |
| Group | **Delta** | **Delta to Omicron** | **Omicron** | ***P* value** | **Delta** | **Delta to Omicron** | **Omicron** | ***P* value** | **Delta** | **Delta to Omicron** | **Omicron** | ***P* value** |
| Fluid and electrolyte disorders | | | | | | | | | | | | |
| no | 61.1% (1,011) | 67.2% (733) | 75.8% (1,326) |  | 75.6% (446) | 82.8% (432) | 83.3% (1,011) |  | 53.1% (565) | 52.9% (301) | 58.9% (315) |  |
| yes | 38.9% (643) | 32.8% (358) | 24.2% (423) | < 0.01 | 24.4% (144) | 17.2% (90) | 16.7% (203) | < 0.01 | 46.9% (499) | 47.1% (268) | 41.1% (220) | 0.06 |
| Blood loss anaemia | | | | | | | | | | | | |
| no | 99.7% (1,649) | 99.6% (1,087) | 99.4% (1,739) |  | 99.3% (586) | 99.6% (520) | 99.3% (1,205) |  | 99.9% (1,063) | 99.6% (567) | 99.8% (534) |  |
| yes | 0.3% (5) | 0.4% (4) | 0.6% (10) | 0.46 | 0.7% (4) | 0.4% (2) | 0.7% (9) | 0.69 | 0.1% (1) | 0.4% (2) | 0.2% (1) | 0.51 |
| Deficiency anaemia | | | | | | | | | | | | |
| no | 97.6% (1,614) | 97.4% (1,063) | 98.0% (1,714) |  | 96.6% (570) | 97.1% (507) | 97.8% (1,187) |  | 98.1% (1,044) | 97.7% (556) | 98.5% (527) |  |
| yes | 2.4% (40) | 2.6% (28) | 2.0% (35) | 0.56 | 3.4% (20) | 2.9% (15) | 2.2% (27) | 0.33 | 1.9% (20) | 2.3% (13) | 1.5% (8) | 0.63 |
| Alcohol abuse | | | | | | | | | | | | |
| no | 97.6% (1,614) | 97.7% (1,066) | 98.1% (1,716) |  | 96.8% (571) | 97.3% (508) | 98.4% (1,195) |  | 98.0% (1,043) | 98.1% (558) | 97.4% (521) |  |
| yes | 2.4% (40) | 2.3% (25) | 1.9% (33) | 0.55 | 3.2% (19) | 2.7% (14) | 1.6% (19) | 0.06 | 2.0% (21) | 1.9% (11) | 2.6% (14) | 0.66 |
| Drug abuse | | | | | | | | | | | | |
| no | 99.6% (1,648) | 99.3% (1,083) | 99.3% (1,737) |  | 99.7% (588) | 98.5% (514) | 99.2% (1,204) |  | 99.6% (1,060) | 100.0% (569) | 99.6% (533) |  |
| yes | 0.4% (6) | 0.7% (8) | 0.7% (12) | 0.34 | 0.3% (2) | 1.5% (8) | 0.8% (10) | 0.10 | 0.4% (4) | 0.0% (0) | 0.4% (2) | 0.34 |
| Psychoses | | | | | | | | | | | | |
| no | 99.3% (1,642) | 99.6% (1,087) | 99.4% (1,738) |  | 99.5% (587) | 99.6% (520) | 99.5% (1,208) |  | 99.2% (1,055) | 99.6% (567) | 99.1% (530) |  |
| yes | 0.7% (12) | 0.4% (4) | 0.6% (11) | 0.48 | 0.5% (3) | 0.4% (2) | 0.5% (6) | 0.94 | 0.8% (9) | 0.4% (2) | 0.9% (5) | 0.45 |
| Depression | | | | | | | | | | | | |
| no | 95.6% (1,581) | 96.0% (1,047) | 97.4% (1,704) |  | 95.1% (561) | 96.4% (503) | 98.2% (1,192) |  | 95.9% (1,020) | 95.6% (544) | 95.7% (512) |  |
| yes | 4.4% (73) | 4.0% (44) | 2.6% (45) | 0.01 | 4.9% (29) | 3.6% (19) | 1.8% (22) | < 0.01 | 4.1% (44) | 4.4% (25) | 4.3% (23) | 0.97 |

SARI = severe acute respiratory infection
